# PReCePT: Equity in antenatal MgSO_4_ treatment

**DOI:** 10.64898/2025.12.16.25341898

**Authors:** Hannah B Edwards, David Odd, Cheryl McQuire, Frank de Vocht, Karen Luyt

## Abstract

This protocol describes the rationale, plan, and methodology for a study investigating sociodemographic inequities in use of antenatal magnesium sulphate (MgSO4) in England, for neuroprotection of the preterm infant. It is a secondary data analysis study using routinely collected healthcare data from the National Neonatal Research Database. The focus of our analysis is to identify whether any inequities in treatment changed from before to after implementation of *PReCePT*, a national Quality Improvement programme to increase antenatal MgSO4 use in England. This work is motivated by, and in the context of, wider evidence of persistent inequities in maternal and perinatal care in England, and evidence that QI programmes, even those that successfully improve overall levels of care, do not necessarily address – any may even worsen – inequities in care.

## Introduction

This study is part of the PReCePT Devolved Nations study, and readers are referred to the original protocol for details of the wider study.

Preterm birth is the leading cause of brain injury and cerebral palsy (CP).(1) Around 1.5% of births are very preterm at less than 30 weeks gestational age (GA).(2) Progress in perinatal care has led to greater survival rates in infants born preterm.(3) However, while 90% of very preterm infants survive beyond the postpartum period, approximately 35% subsequently develop neurodisabilities, including CP, blindness, deafness and cognitive impairment.(4)

Antenatal magnesium sulphate (MgSO_4_) given to women at risk of preterm birth reduces the risk of CP in their child by 30% (relative risk (RR) 0.68; 95% Confidence Interval (CI) 0.54 to 0.87).(5) It has been estimated that around 200 cases of CP in England could be avoided by consistent administration of MgSO_4_ during labour each year.(6) At under 33 weeks gestation, the number needed to treat (NNT) to prevent one case of CP is 42 (95% CI 26 to 187), and this reduces to a NNT of 37 in births under 30 weeks gestation.(6) The impact of CP is significant and lifelong for individuals, families and health services.(7) Administration of MgSO_4_ can be highly cost-effective with an estimated £1M lifetime societal savings per case of CP avoided.(8, 9)

Since 2015 the National Institute for Health and Care Excellence (NICE) has recommended administration of MgSO_4_ in very preterm deliveries (under 30 weeks GA) as a core part of maternity care.(10) Failing to comply with this guideline is considered sub-optimal care. However, until recently, uptake of MgSO_4_ in the UK has been low. For infants below 30 weeks GA, uptake was 9%, 24% and 38% for 2012, 2013, and 2014 in the UK respectively compared to 46%, 51% and 56% for the international network.(11) By 2017 UK uptake was at 64%. High regional variation (range 49% to 78%) indicated inequalities in perinatal care.(12) This slow uptake, as was also seen in the case of antenatal steroids, which took decades to become embedded in clinical practice, is associated with high health and economic costs. Investment in initiatives to accelerate uptake is likely to benefit patients and families.

The PReCePT (Preventing Cerebral Palsy in Pre-Term labour) Quality Improvement (QI) toolkit and implementation guide was developed to improve maternity staff awareness and thereby increase use of MgSO_4_. Following a positive pilot study in 2018,(13) this was scaled-up and rolled-out across England as the National PReCePT Programme (NPP). The aim was to increase MgSO_4_ use in England to 85% by 2020. Units received the PReCePT QI toolkit, comprising of an implementation guide and additional resources (pre-term labour proforma, staff training presentations, parent information leaflet, posters for the unit, learning log)(14) Each unit had a lead midwife as ‘PReCePT champion’, and the NPP provided up to 90 hours funded backfill for this role. Coaching and support was available through the 15 Academic Health Science Networks (AHSNs), via a regional clinical lead (obstetrician and/or neonatologist).

We evaluated the effectiveness and cost-effectiveness of the NPP in increasing MgSO_4_ uptake in England. The first evaluation covered the first 12 months of the programme and found average MgSO_4_ uptake increased by 6.3 percentage points (95% CI 2.6 to 10.0 percentage points) to 83.1% post-implementation (adjusted for unit size, maternal, baby, and maternity unit factors, time trends, and AHSN region). The health gains and cost-savings associated with the NPP generated a net monetary benefit of £866 per preterm baby. The probability of the NPP being cost-effective was greater than 95%.(15) The second evaluation (the Devolved Nations extension study) aims to evaluate the longer-term effectiveness and cost-effectiveness of the programme over the first four years following roll-out, and additionally compare MgSO_4_ use in England with Scotland and Wales. The full study is detailed elsewhere(16) (protocol and SAP also available).

One workstream within the extension study is to evaluate individual-level risk factors for mothers receiving MgSO_4_, to elucidate any inequities in treatment, and changes in equity over time. The protocol outlined here details this workstream specifically. There is evidence from previous work that there are inequities in maternal and perinatal care. For example, the MBRRACE-UK Perinatal Confidential Enquiries reports have consistently found racial differences in care of women who experienced stillbirth or neonatal death, and in maternal death rates (most recent 2024).(17) The Royal College of Obstetrics and Gynaecology National Maternity and Perinatal Audit Inequalities report 2021 found differences by race and level of socio-economic deprivation in risk of caesarean birth, blood loss after childbirth, baby being small for gestational age (SGA), Apgar score, admission to a neonatal unit, and breastfeeding.(18) A UK national cohort study in 2021 found socio-economic and ethic disparities in stillbirths, preterm births, and births with fetal growth restriction.(19) Another study from England in 2020 found socio-economic, ethic, and maternal age disparities in maternal mortality, with ethnic disparities worsening and socio-economic disparities improving over time.(20) As well as these socio-demographic variations, the latest annual report from the National Neonatal Audit Programme(21) indicates geographical (regional) disparities across England in many outcome metrics including neonatal mortality, brain injury, bronchopulmonary dysplasia (BPD), necrotising enterocolitis (NEC), antenatal steroid delivery, MgSO_4_ delivery, deferred cord clamping, normal temperature, and breastmilk feeding.

In this study we aim to explore disparities in maternal receipt of MgSO_4_ specifically.

### Research Aim

Identify any disparities in pregnant people (hereafter referred to as mothers) receiving antenatal MgSO_4_ in England, and any changes in disparities over time.

### Objectives

#### Primary

1. Identify sociodemographic disparities in receipt of MgSO_4_ by maternal age, ethnicity, and area deprivation.
2. Identify geographic disparities in receipt of MgSO_4_ by North versus South of England.
3. Identify any changes in disparities from before to after the launch of the National PReCePT Programme (NPP).

### Hypotheses

1. Sociodemographic disparities in receipt of MgSO_4_ will have reduced post-NPP compared to pre-NPP.
2. Geographic disparities in receipt of antenatal MgSO_4_ will have reduced post-NPP compared to pre-NPP.

### Study design

This is a cohort study using routinely collected, longitudinal observational patient data.

## Methods: participants, intervention, and outcomes

### Study setting

Maternity and neonatal units in England.

### Study duration

We anticipate the data time frame to include babies born between January 2014 and October 2024.

### Eligibility criteria

#### Inclusion criteria

- Mothers with a baby born prematurely between 24+0 and 29+6 weeks gestation
- The pregnancy was booked in an English hospital and baby born in England
- Singletons and first infant of multiple births

#### Exclusion criteria

- Mothers with babies born under 24 weeks, or born at 30 weeks or more. This is because the NICE guidance on antenatal MgSO_4_ recommends treatment for babies under 30 weeks gestation. (They recommend ‘consideration’ of treatment for babies 30-34 weeks gestation; this older group is not included in this study.)
- Pregnancy not booked in England or baby not born in England. This is because study approvals cover England only.
- Mother or baby missing unique identifier code in the data.

#### Intervention

The intervention being evaluated is the National PReCePT Programme, in terms of its potential effect on inequities in receipt of antenatal MgSO_4_.

#### Outcomes

The primary outcome is receipt of antenatal MgSO_4_ (yes versus no) at the individual (mother) level.

#### Sample size

Data from all mothers meeting the inclusion criteria above are included in this study. We anticipate 155 English maternity/neonatal units to be included. Data from the original study suggests that we can anticipate around 3000 per year age 24+0 to 29+6 weeks gestation (and around 6700 per year age 30+0 to 33+6 weeks gestation).

#### Recruitment

Recruitment is not applicable as this study uses only routinely collected healthcare data.

## Methods: data collection, management, and analysis

### Data collection methods

Routine pseudonymised patient-level data from the UK National Neonatal Research Database (NNRD) is used. Consent to use data for each unit is obtained centrally by the Neonatal Data Analysis Unit (NDAU), which manages the NNRD.

### Data management

Storage of all data will comply with the General Data Protection Regulation (2018) and University of Bristol’s data protection policies. Storage will be on secure University computer systems within a Safe Haven folder. The Chief Investigator acts as data custodian.

### Analysis

All quantitative data will be analysed using STATA version 18 (Stata Statistical Software: Release 18. College Station, TX: StataCorp LLC).

#### Variables

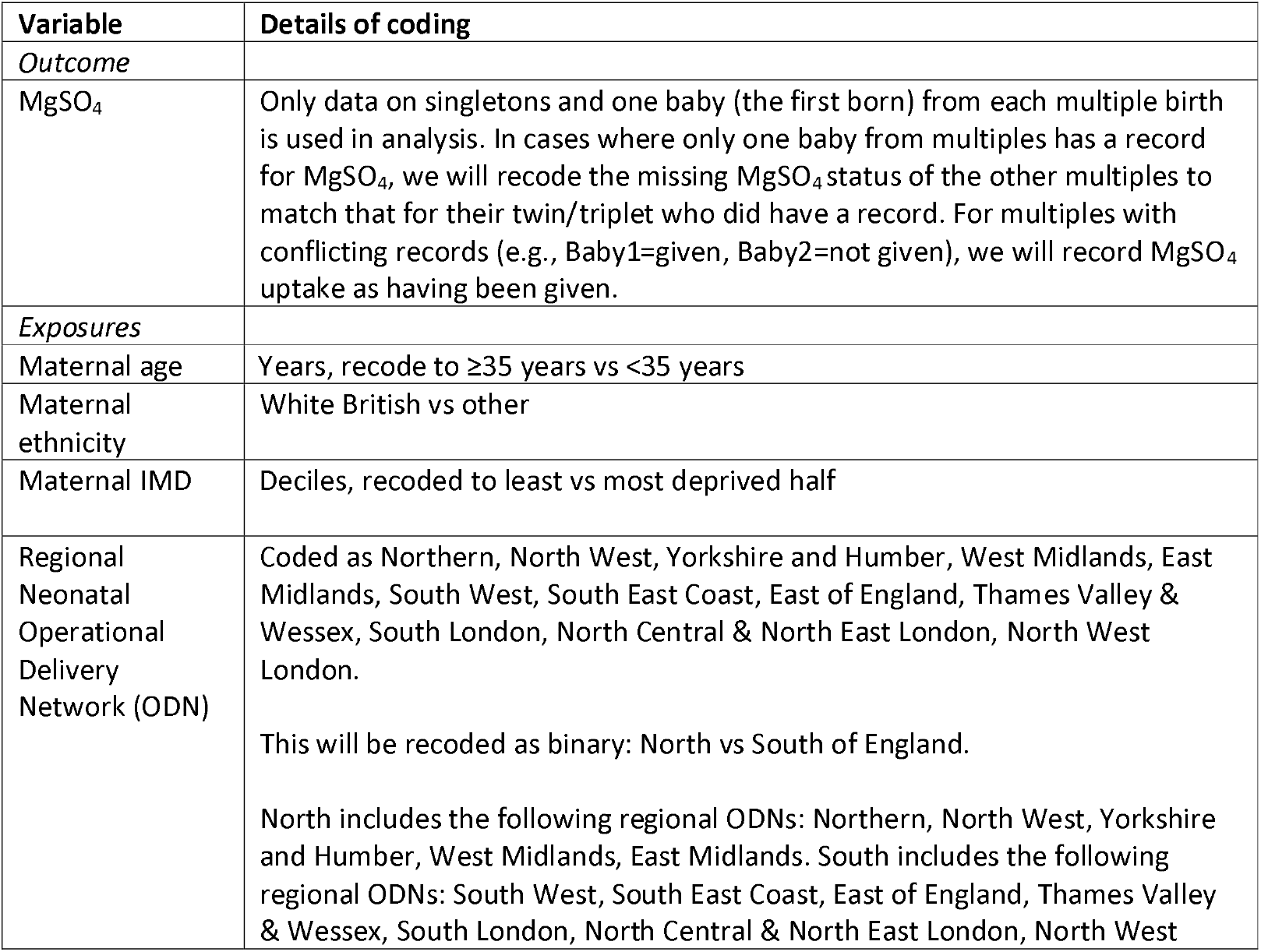
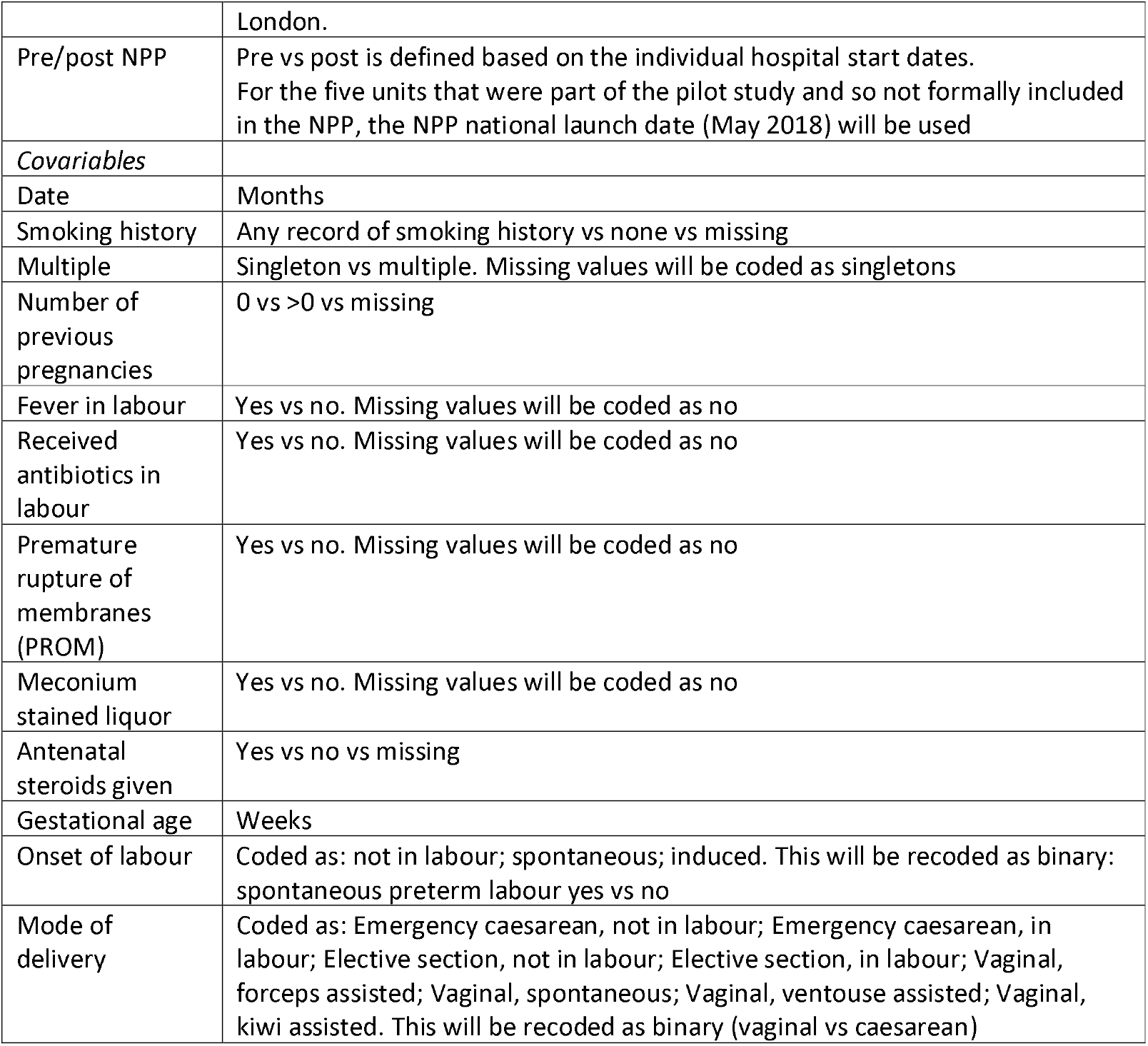

| Variable | Details of coding |
| --- | --- |
| *Outcome* |  |
| MgSO<sub>4</sub> | Only data on singletons and one baby (the first born) from each multiple birth is used in analysis. In cases where only one baby from multiples has a record for MgSO<sub>4</sub>, we will recode the missing MgSO<sub>4</sub> status of the other multiples to match that for their twin/triplet who did have a record. For multiples with conflicting records (e.g., Baby1=given, Baby2=not given), we will record MgSO<sub>4</sub> uptake as having been given. |
| *Exposures* |  |
| Maternal age | Years, recode to ≥35 years vs <35 years |
| Maternal ethnicity | White British vs other |
| Maternal IMD | Deciles, recoded to least vs most deprived half |
| Regional Neonatal Operational Delivery Network (ODN) | Coded as Northern, North West, Yorkshire and Humber, West Midlands, East Midlands, South West, South East Coast, East of England, Thames Valley & Wessex, South London, North Central & North East London, North West London.<br>This will be recoded as binary: North vs South of England.<br>North includes the following regional ODNs: Northern, North West, Yorkshire and Humber, West Midlands, East Midlands. South includes the following regional ODNs: South West, South East Coast, East of England, Thames Valley & Wessex, South London, North Central & North East London, North West |
|  | London. |
| Pre/post NPP | Pre vs post is defined based on the individual hospital start dates.<br>For the five units that were part of the pilot study and so not formally included in the NPP, the NPP national launch date (May 2018) will be used |
| *Covariables* |  |
| Date | Months |
| Smoking history | Any record of smoking history vs none vs missing |
| Multiple | Singleton vs multiple. Missing values will be coded as singletons |
| Number of previous pregnancies | 0 vs >0 vs missing |
| Fever in labour | Yes vs no. Missing values will be coded as no |
| Received antibiotics in labour | Yes vs no. Missing values will be coded as no |
| Premature rupture of membranes (PROM) | Yes vs no. Missing values will be coded as no |
| Meconium stained liquor | Yes vs no. Missing values will be coded as no |
| Antenatal steroids given | Yes vs no vs missing |
| Gestational age | Weeks |
| Onset of labour | Coded as: not in labour; spontaneous; induced. This will be recoded as binary: spontaneous preterm labour yes vs no |
| Mode of delivery | Coded as: Emergency caesarean, not in labour; Emergency caesarean, in labour; Elective section, not in labour; Elective section, in labour; Vaginal, forceps assisted; Vaginal, spontaneous; Vaginal, ventouse assisted; Vaginal, kiwi assisted. This will be recoded as binary (vaginal vs caesarean) |

#### Descriptive analysis

The distribution of demographic, geographic, and clinical factors will be described for pre and post-NPP. Continuous variables will be summarised using means and standard deviations (SD) (or medians and interquartile ranges (IQR) if the distribution is highly skewed), and categorical data will be summarised as numbers and percentages. The number and percentage of mothers receiving MgSO_4_ by year will be presented. For consistency with nationally reported data (National Neonatal Audit Programme (NNAP) reporting), MgSO_4_ uptake is defined as the number of mothers recorded as having received MgSO_4_ at a Unit divided by the total number of eligible mothers at that Unit, excluding missing values from the denominator. This will be expressed as a percentage.

#### Main analysis

Logistic regression models will be developed for each characteristic, generating an odds ratio (OR) representing odds of receiving MgSO_4_ if a mother was exposed, divided by odds of receiving MgSO_4_ if a mother was not exposed. ORs below 1.0 indicate that the characteristic is associated with lower odds of receiving MgSO_4_. ORs above 1.0 indicate that the characteristic is associated with higher odds of receiving MgSO_4_. ORs will be presented with 95% confidence intervals (95% CI) and p-values. If the 95% CI excludes 1.0, that will be interpreted as evidence for an association between the characteristic and receipt of MgSO_4_.

Unadjusted and adjusted multivariable models will be developed for each characteristic and the outcome of receiving MgSO_4_. To evaluate changes in equity from pre- to post-NPP, both unadjusted and adjusted models will include a binary variable for pre-versus post-NPP, and interaction between this and the characteristic of interest. Where there is evidence of statistically significant interaction, the coefficients for pre- and post-NPP will be presented separately.

The variables included in the adjusted models have been decided *a priori* based on the minimally adjusted set from Directed Acyclic Graphs (DAGs)(22), developed by the research team, using the DAGitty software programme (https://www.dagitty.net/)(23). For each exposure of interest, we will show two different adjusted models, one showing the *total effects* of that exposure, and one showing the direct effects of that exposure. Separate confounder sets have been considered, and developed, for each exposure of interest and to estimate both total and *direct effects*. These adjustment sets for each exposure of interest, together with their DAGs, are included as an appendix in this protocol. Full coding of each model within the DAGitty programme is available from the research team.

Models will be multi-level, with maternity unit as a higher level variable; to account for clustering by maternity unit. Here the individual-level data is the first level (fixed effects), nested in maternity unit as the second level (random effects). To account for the underlying time trend, time (in months) will also be included in the models as a first level variable.

#### Sensitivity analysis

As a sensitivity analysis will re-run the models excluding mothers with a record of pregnancy hypertension. This is because in theory (although uncommonly in practice) MgSO_4_ can be given to a mother with hypertension as a prophylactic against seizures. As this is in theory a distinct group from mothers given antenatal MgSO_4_ for neuroprotection of the infant, we will check that main results are robust if we exclude them.

## Data Availability

NNRD data dictionaries are publicly available at https://digital.nhs.uk/data-and-information/information-standards/governance/latest-activity/standards-and-collections/dapb1595-neonatal-data-set/. NNRD data is accessible via formal application.

## Glossary of Abbreviations

AHSN: Academic Health Science Network
ARC: Applied Research Collaboration
CI: Confidence Interval
CP: Cerebral Palsy
FREC: Faculty Research Ethics Committee
GA: Gestational Age
GCP: Good Clinical Practice
HRA: Health Research Authority
MgSO_4_: Magnesium Sulphate
NDAU: Neonatal Data Analysis Unit
NHS: R&D National Health Service Research & Development
NICE: National Institute for Health and Care Excellence
NIHR: National Institute for Health and Care Research
NNAP: National Neonatal Audit programme
NNRD: National Neonatal Research Database
NPP: National PReCePT Programme
ODN: Operational Delivery Network
OR: Odds Ratio
PreCePT: Prevention of Cerebral Palsy in pre-term labour
RCT: Randomised Control Trial
SAP: Statistical Analysis Plan
SOP: Standard Operating Procedure
VON: Vermont and Oxford network
WEAHSN: West of England Academic Health Science Network

## Ethics, Regulatory issues, and dissemination

### Research Ethics

No informed consent is sought from individual women delivering in maternity units for their participation in this study, as health care provided is according to current practice, following the clinical practice NICE Preterm labour and birth guideline (NG25) [7], at the discretion of the obstetrician/midwife. Only routinely collected pseudonymised data will be used, which are not subject to NHS ethical review. The wider PReCePT Devolved Nations study, of which this is a part, has received research permissions and approvals from the HRA, and Sponsor faculty approval, as detailed on the covering page.

### Indemnity

For the wider PReCePT Devolved Nations study, the University of Bristol has arranged Public Liability insurance to cover the legal liability of the University as Research Sponsor in the eventuality of harm to a research participant arising from management of the research by the University. This does not in any way affect an NHS Trust’s responsibility for any clinical negligence on the part of its staff (including the Trust’s responsibility for University of Bristol employees acting in connection with their NHS honorary appointments).

### Study Sponsorship

The research is sponsored by the University of Bristol and is organised and funded by The AHSN Network and The National Institute for Health Research Applied Research Collaboration (NIHR ARC West). This extension is funded by The Heath Foundation and ARC West. This study has been reviewed by University of Bristol Health Sciences Faculty Research Ethics Committee (FREC).

### Patient and Public Involvement and Engagement (PPIE)

The original PReCePT1 intervention was developed using a co-design and co-production approach, developing partnerships with BLISS, a support organisation for mothers experiencing pre-term births, and two mothers who had experienced pre-term births. These two mothers are part of the steering group for the wider PReCePT programme evaluation, enabling the project to benefit from the knowledge and experiences they have developed over the life of the PReCePT implementation.

### Dissemination

All outputs from this research will be written and presented according to the NIHR ARC West publication policy. We plan to publish and share the results of study in relevant professional network publications, including peer reviewed journals. We will present the work by disseminating it to a range of networks for both clinical and academic audiences. We aim to achieve regional and national impact through presentations at national and international scientific conferences and workshops.

## Appendices Appendix 1

Adjustment sets for models

Appendix 1: Adjustment sets for models
| Exposures of interest | Outcome | Model 1: Unadjusted | Model 2: TOTAL EFFECTS | Model 3: DIRECT EFFECTS |
| --- | --- | --- | --- | --- |
| Maternal age (≥35 vs <35) | MgSO<sub>4</sub> | Pre/Post NPP<br>Interaction between exposure and Pre/Post NPP | *Model 1 +*<br>IMD<br>Ethnicity<br>Region<br>+date<br>+clustering on unit | *Model 2 +*<br>GA<br>Multiple<br>Prev. pregnancies<br>Smoking |
| IMD (high deprivation vs low deprivation) | MgSO<sub>4</sub> | Pre/Post NPP<br>Interaction between exposure and Pre/Post NPP | *Model 1 +*<br>Ethnicity<br>Region<br>+date<br>+clustering on unit | *Model 2 +*<br>GA<br>Delivery mode<br>Fever in labour<br>Maternal age<br>Multiple<br>Onset of labour<br>Prev. pregnancies<br>Prom<br>Smoking |
| Ethnicity (white vs non-white) | MgSO<sub>4</sub> | Pre/Post NPP<br>Interaction between exposure and Pre/Post NPP | *Model 1 +*<br>No others<br>*("No open biasing paths. No adjustment is necessary to estimate the total effect of ethnicity on MgSO<sub>4</sub>")*<br>+date<br>+clustering on unit | *Model 2 +*<br>IMD<br>Maternal age<br>Prev. pregnancies<br>Region<br>Smoking |
| Region (north vs south) | MgSO<sub>4</sub> | Pre/Post NPP<br>Interaction between exposure and Pre/Post NPP | *Model 1 +*<br>Ethnicity<br>+date<br>+clustering on unit | *Model 2 +*<br>GA<br>IMD<br>Delivery mode<br>Fever in labour<br>Maternal age<br>Multiple<br>Onset of labour<br>Prev. pregnancies<br>Prom<br>Smoking |

## Appendix 2

Directed Acyclic Graphs for models

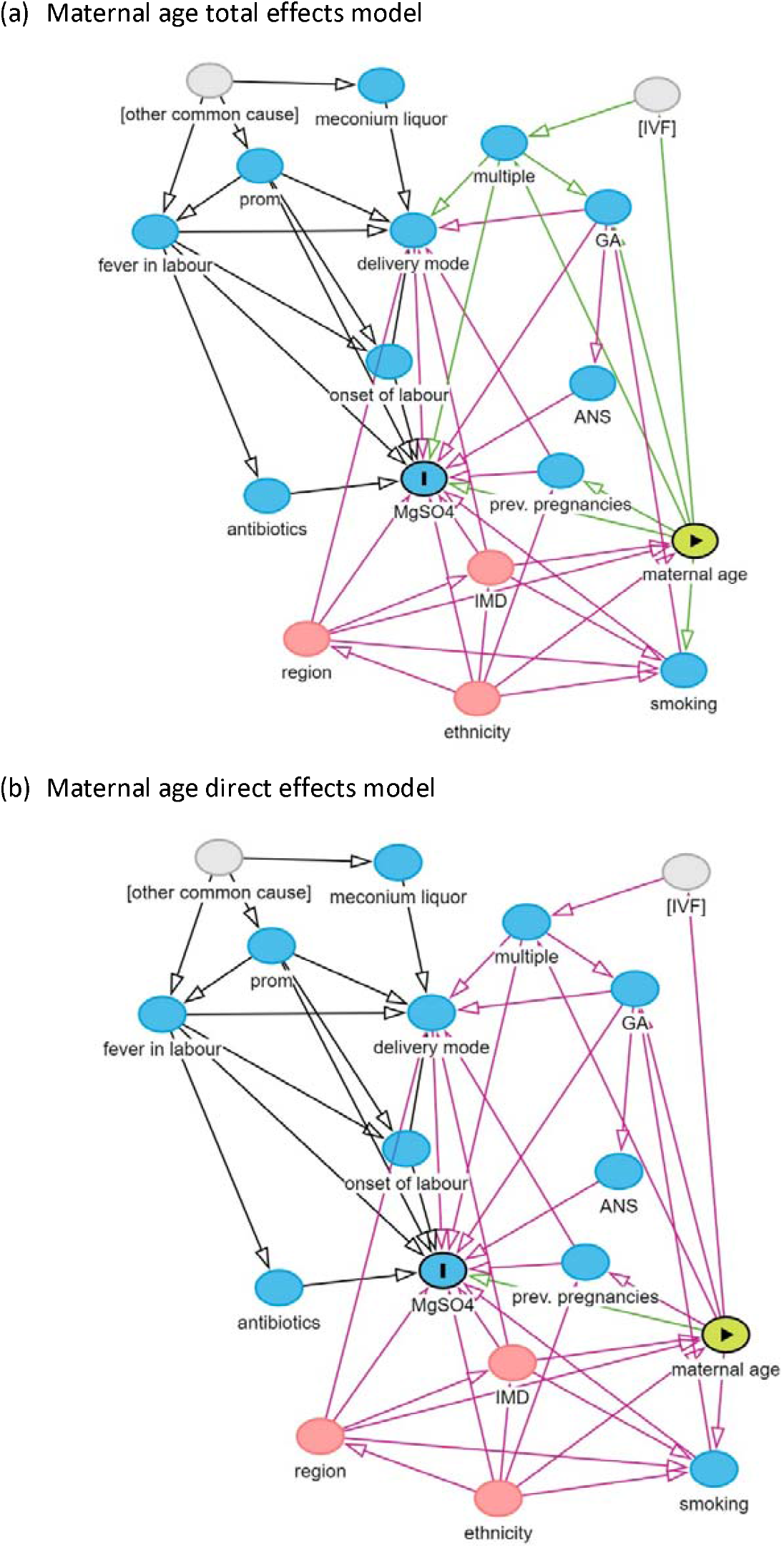
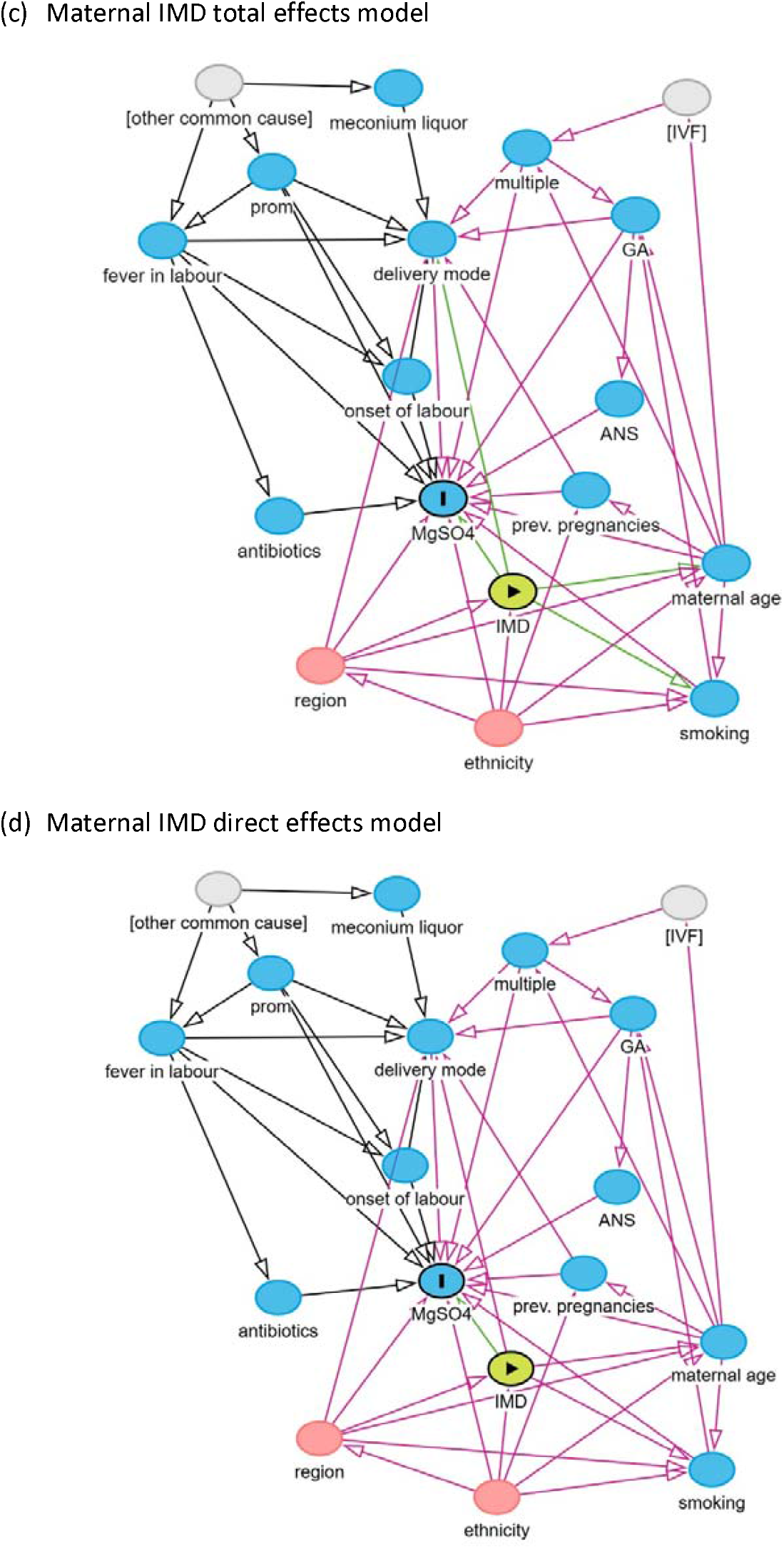
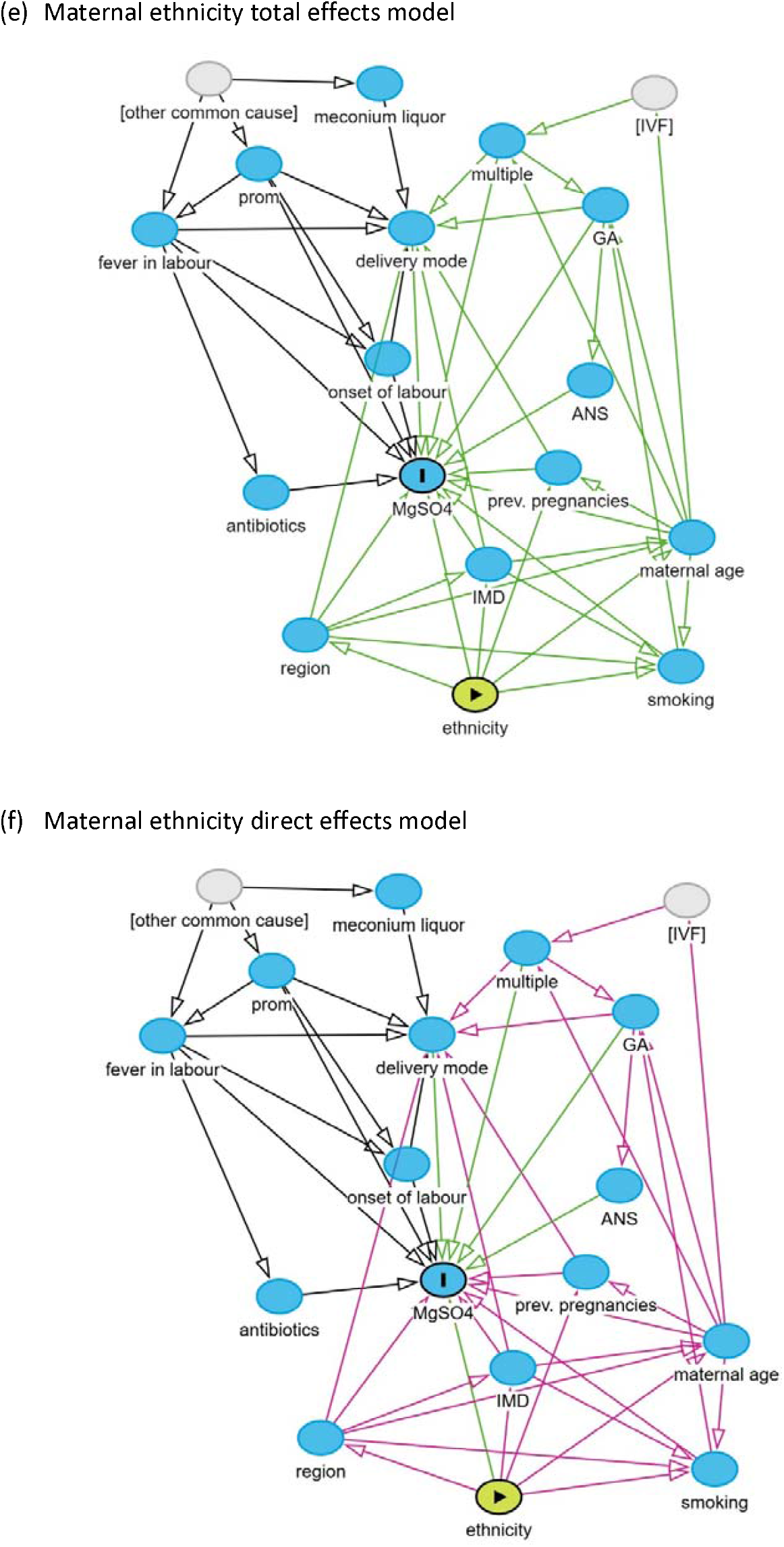
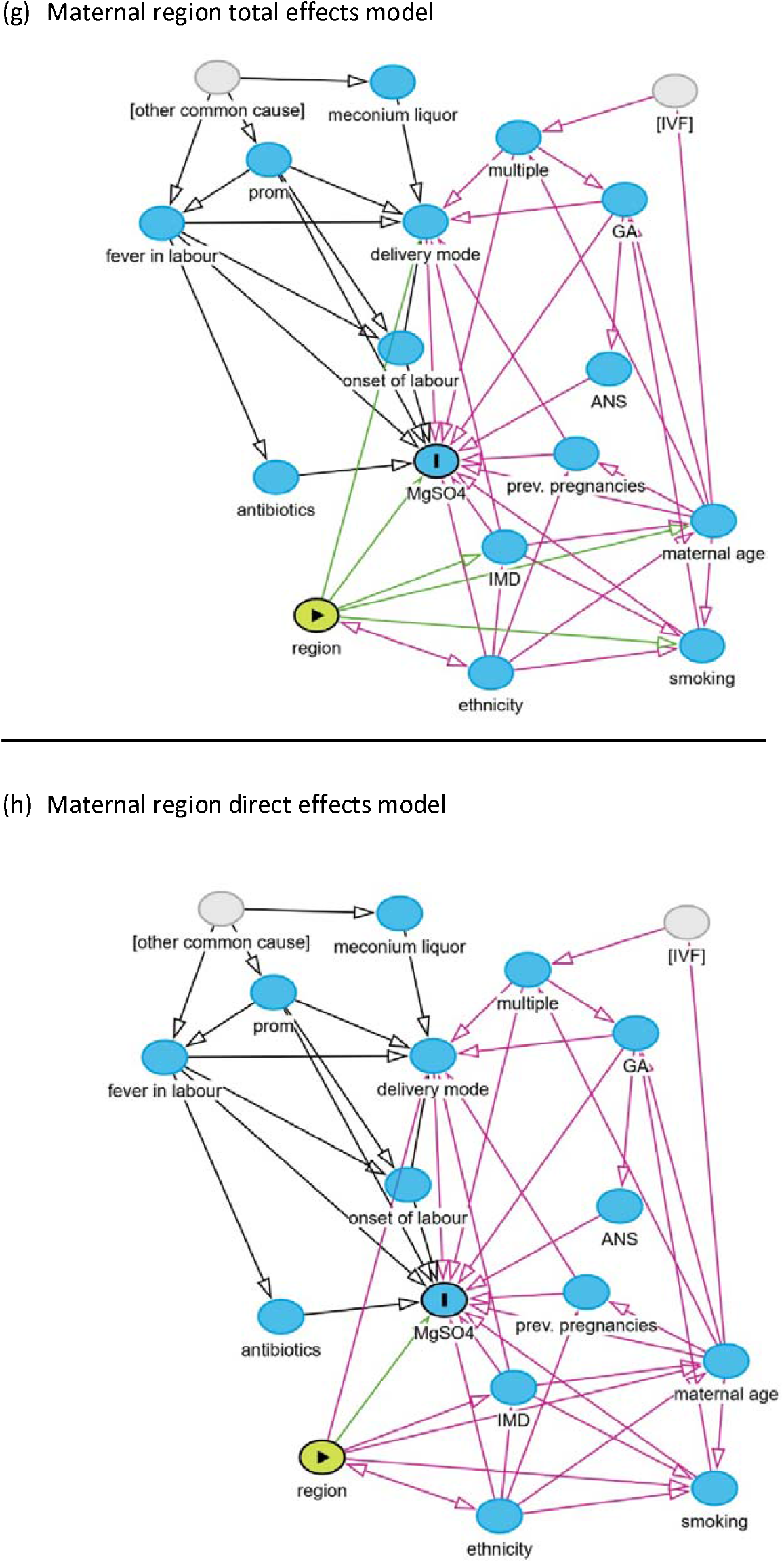

## Appendix 3

DAG models key

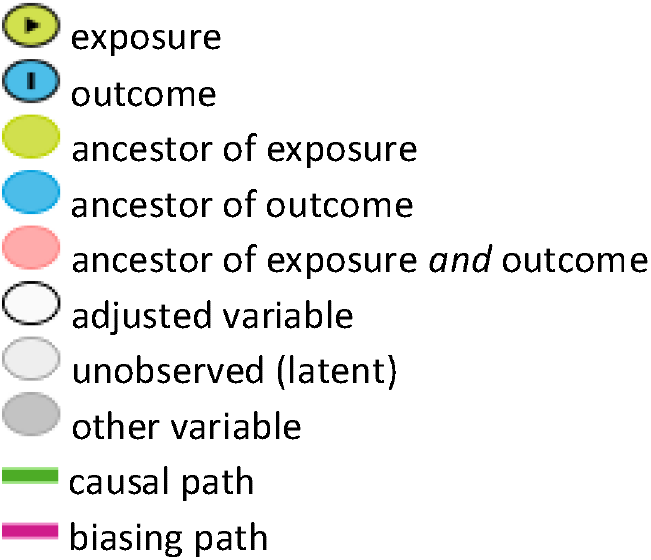

